# Mobility Function and Aperiodic Electrocortical Activity in Younger and Older Adults

**DOI:** 10.1101/2025.08.31.25334794

**Authors:** Charlotte R. DeVol, Chang Liu, Jacob Salminen, Erika M. Pliner, Arkaprava Roy, Chris J. Hass, David J. Clark, Todd M. Manini, Rachael D. Seidler, Daniel P. Ferris

## Abstract

Mobility declines with age to the extent that walking speed is often considered a vital sign. Identifying neurological mechanisms behind this decline would aid with early identification and intervention. Electroencephalography (EEG) metrics may provide insight into neural factors contributing to mobility decline with aging. Recent research has shown a differentiation in aperiodic EEG metrics (exponent and offset) across age groups, cognitive abilities, and populations with neurological injury. However, it is unknown if aperiodic EEG differs between mobility tasks or brain regions. The purpose of this study was to 1) compare aperiodic EEG in older and younger adults at rest and while walking and 2) determine if oscillatory and aperiodic EEG in sensorimotor brain regions are predictors of a lower walking speed, regardless of age and other demographic factors. We analyzed EEG collected while participants were sitting at rest and walking on a treadmill in 31 younger adults (age: 24 ± 4, mean ± s.d.) and 59 older adults (age: 74 ± 6). We found that older adults had lower aperiodic exponent and offset at rest and during walking, but only a subset of brain regions showed age group differences. Using machine learning methods, we found that right sensorimotor alpha and left sensorimotor offset and beta had the largest effect on individualized walking speed, after the demographics of age, waist circumference, and sex. These results suggest aperiodic and oscillatory EEG in specific brain areas may give additional insights into brain health.

## I. INTRODUCTION

Many older adults experience a decline in mobility with age. This decline often includes walking at slower speeds, increased movement variability, and greater reliance on cognitive over automatic control [1], [2]. With aging comes changes in the recruitment of different brain areas, which can be a contributing factor to declining mobility [3], [4], [5]. Specifically, older adults rely on more widespread recruitment of the basal ganglia, cerebellum, frontal and parietal cortices [3], [6], [7]. Understanding how changes in brain health relate to a decline in mobility with aging is beneficial for designing timely and effective interventions. Quantifying neural dynamics while walking may give additional insight into the specific mechanisms driving mobility decline with age, such as decreased walking speed.

Developments in mobile brain imaging with electroencephalography (EEG) allow for direct, non-invasive measurement of electrocortical activity during whole-body tasks [8], [9], [10]. Mobile EEG analysis primarily focuses on measuring changes in oscillatory (i.e. periodic) EEG. For example, prior work has observed changes in alpha (8-13 Hz) and beta (13-30 Hz) spectral power when participants walk on a balance beam, on uneven grass terrain, and at different speeds [11], [12], [13], [14]. We have also observed that compared to younger adults, older adults adjusted theta power (4–7 Hz) different in some brain regions when increasing gait speed [14]. The EEG power in specific frequency bands represents dynamic communication across populations of neurons [15]. There are also broadband shifts in EEG that affect all frequencies [16]. Failure to separate the oscillatory from the broadband EEG, also known as aperiodic EEG, can result in inaccurate interpretation of oscillatory EEG [17], [18]. However, aperiodic EEG remains less studied, especially during whole-body movement.

For many years aperiodic EEG, also known as neural noise or 1/f behavior, was thought to be a result of filtering of the cortical signal as it passed from the brain to the EEG electrode on the scalp. While filtering may occur, more recent studies have shown that shifts in broadband neural activity can occur with changes in population-level synchrony of neural networks [19], [20], [21], the release of neurotransmitters for excitation and inhibition in the brain [22], [23], and changes in neuronal firing rates [20], [21].

Evidence for aperiodic EEG to represent underlying brain health is present in studies across clinical populations. Aperiodic EEG has shown to be lower in older adults [17], [24], [25], [26], differ in adolescents with attention deficit hyperactivity disorder compared to their peers [27], be a strong predictor of schizophrenia [28], and differ in several other clinical populations [29]. Aperiodic EEG is also related to the variability of visual and cognitive processing speed and performance [30], [31], [32] and explains age-related declines in visual working memory [16]. Decline in cognition with aging, especially working memory, is also largely related to declines in mobility, such as walking speed [33], [34], [35]. However, it remains unclear how aperiodic components of EEG signals relate to decline in mobility with aging. Quantifying aperiodic EEG in younger and older adults and its relationship with mobility may give additional insight beyond oscillatory EEG.

The purpose of this study was to 1) compare the aperiodic EEG across brain regions in older and younger adults at rest and while walking, and 2) determine if oscillatory and aperiodic EEG in the sensorimotor areas are indicators of participant mobility, after controlling for age and other biological variables. We hypothesized that older adults would have lower aperiodic offsets and exponents than younger adults during both rest and walking conditions. We based this hypothesis on prior work that found aperiodic EEG differences across age groups persisted across memory tasks [16]. We tested this hypothesis by comparing differences in aperiodic exponent and offset between younger and older adults in different brain regions during rest and walking at an individualized treadmill speed. We further hypothesized that aperiodic EEG would be a larger predictor of individualized walking speed than oscillatory EEG after controlling for differences in age, waist circumference as a measure of body fat, sex, and other EEG metrics. This hypothesis was based on evidence that aperiodic EEG represents, at least in part, underlying neural processes such as excitation, inhibition and neuronal firing rates [20], [21], [22]. We developed a Bayesian Additive Regression Trees (BART) model to test this hypothesis in the sensorimotor brain areas [36]. BART can model nonlinear relationships and handle missing data using Bayesian methods, making it useful for datasets from human subjects [37]. We quantified the relationship between each model predictor (age, waist circumference, sex, and aperiodic and oscillatory EEG metrics in each of the left and right sensorimotor regions) on participants’ individualized walking speed using Accumulated Local Effects (ALE) plots [38]. From the ALE plots, we calculated the net effect of each predictor on walking speed as an indicator of which predictors have the largest effect on speed relative to other model inputs. The results from this study will improve our understanding of how to interpret aperiodic EEG data and whether it should be considered in future studies on mobility and aging.

## II. METHODS

The data reported here are from a larger study investigating walking and mobility decline in older adults across brain imaging modalities (NIH U01AG061389) [39]. In this analysis, we calculated cross-sectional aperiodic EEG data collected on younger and older adults. The experimental protocol and findings from the oscillatory EEG data have been reported elsewhere [10], [14], [40], [41]. Here we provide a summary of overlapping methods included in prior work.

### A. Participants

We recruited 35 younger adults (aged 20 to 40 years) and 96 older adults (aged 65 and above) to participate in the study. To be included, potential participants had to be able to complete a 400-meter walk test in under 15 minutes without assistance. Potential participants were excluded if they had a Montreal Cognitive Assessment (MoCA) score below 26 or other existing medical conditions such as severe cardiovascular disorders, terminal illness, visual impairment that cannot be corrected, and implants with contraindications to Magnetic Resonance Imaging (MRI). Full inclusion and exclusion criteria have been described previously [39]. A total of 28 participants were excluded from data analysis because of missing an MRI scan (N = 8), inability to complete walking tasks (N = 17), or other technical issues (N = 3). All participants provided written informed consent before joining the study. This study was approved by the University of Florida Institutional Review Board (IRB 201802227).

### B. Experimental Protocol

Participants walked on a customized slat-belt treadmill for a total of approximately 48 minutes (PPS 70 Bari-Mill, Woodway, Waukesha, WI, USA; 70 cm x 173 cm walking surface) (Fig. 1). They either walked at a subject-specific speed based on preferred walking speed on an uneven terrain condition or at one of four standardized speeds on a flat surface (0.25, 0.5, 0.75, and 1.0 m/s). Each condition was performed twice for three minutes. Subject-specific walking speeds were set at 75% of the slowest speed the participant used when walking overground on uneven terrains, unless they requested a slower treadmill speed. During walking trials, all participants wore a safety harness with sufficient overhead slack to avoid supporting the body unless an actual fall were to occur. Participants also completed a seated rest trial for three minutes. Results reported in this manuscript only include the rest and flat walking at the subject-specific walking speeds to focus on task-specific difference between rest and walking. We also measured waist circumference as an indicator of excess body fat and overall physical health [42], [43]. Waist circumference was measured at the midpoint between highest point of iliac crest and lowest part of costal margin in the mid-axillary line. Two measurements were taken initially and if they varied from each other more than 0.5 cm, then a third measurement was taken. Measurements were averaged for one value per person.

**Fig. 1.**
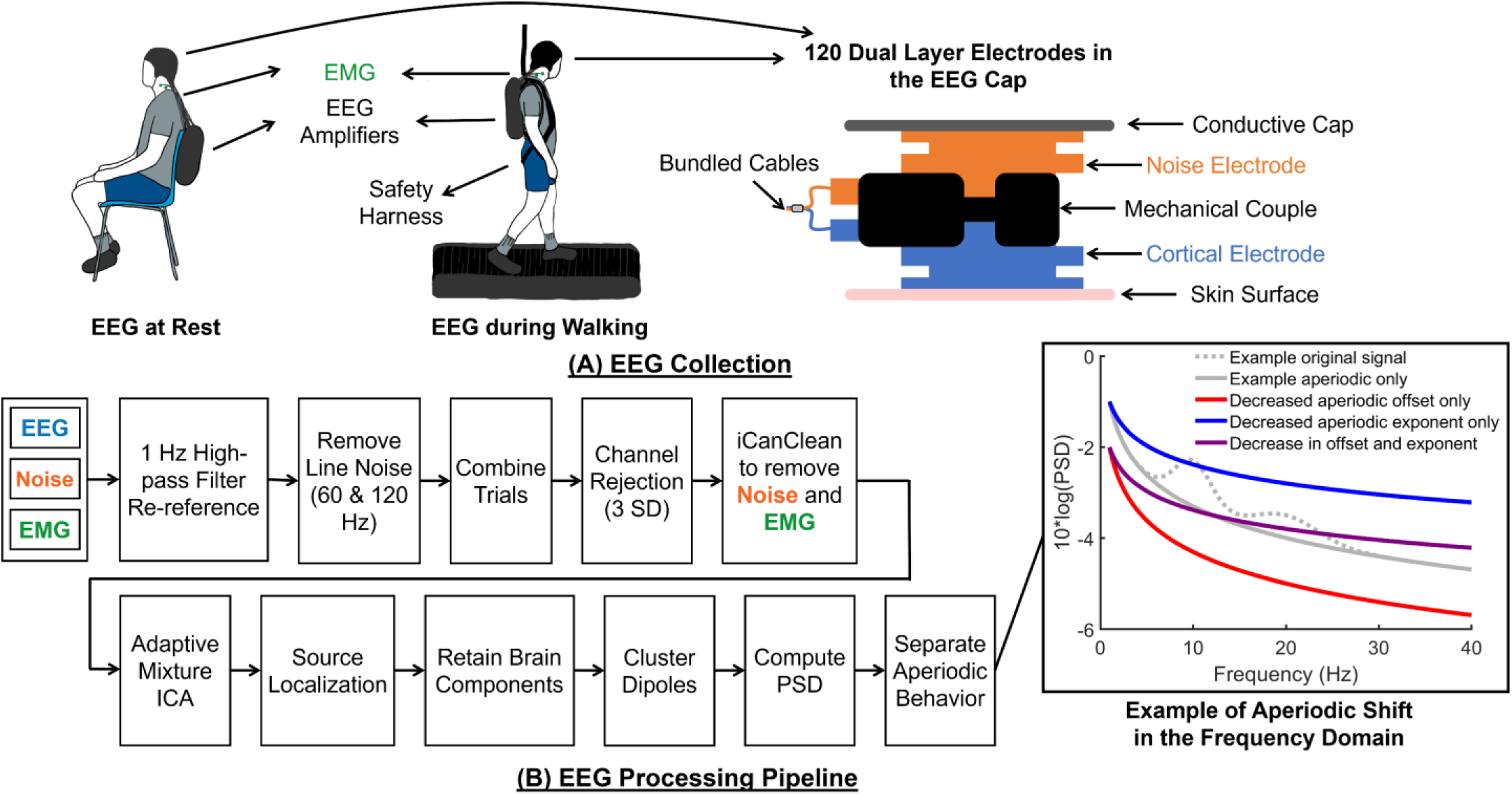
Experimental set-up and data analysis pipeline for electroencephalography (EEG) data collection on older and younger adults at rest and while walking on a treadmill at an individualized speed. (A) Participants complete one 3-minute trial seated at rest and two 3-minute trials walking on a slat-belt treadmill. Participants wore a custom dual-layer EEG cap with 120 recording cortical EEG mechanically coupled to 120 noise EEG. Noise electrodes faced away from the head into a conduction cap to bridge the electrodes. Eight electrodes were repurposed to record electromyography (EMG) of the bilateral sternocleidomastoid and trapezius muscles. (B) Steps for processing EEG data, including noise removal, adaptive mixture independent component analysis (ICA), source localization, identifying brain components, and separating aperiodic and oscillatory EEG. An example of shifts in aperiodic exponent and offset are provided in the far-right graph.

### C. Data Acquisition

To record electrocortical activity during each task, participants wore a custom-made dual-layer EEG cap (ActiCAP snap; Brain Products GmbH, Germany). The electrode layout consisted of 120 electrodes facing the scalp following a 10-05 electrode system, 120 noise electrodes mechanically coupled to the scalp electrodes facing away from the head, and 8 electrodes (originally TP9, P9, PO9, O9, O10, PO10, P10, and TP10) repurposed to record the electromyography (EMG) of the bilateral sternocleidomastoid and trapezius muscles [44], [45]. During cap set-up, the impedance of all scalp electrodes was kept below 15 kΩ and below 5 kΩ for the ground and reference electrodes, Fpz and Cpz, respectively. The EEG cap was covered with a custom-made cap of conductive graphene doped fabric (Eeontex LTT-PI-100, Marktek Inc., USA) to act as an artificial skin surface for the noise electrodes recording motion artifact [46]. The EEG system recorded at 500 Hz with four LiveAMP64 amplifiers (Brain Products GmbH, Germany). We used a 3D structural scanner to digitize the location of electrodes (ST01, Occipital Inc., San Francisco, CA, USA).

All participants participated in a structural MRI scan from a T1-weighted scanner on a separate day than the EEG data collections. The visits were within 30±91 days of each other. Imaging parameters for the MRI scanner included: repetition time = 2000 ms, echo time = 2.99 ms, flip angle = 8°, voxel resolution = 0.8 mm^3^, and field of view = 256 × 256 × 167 mm^2^ using a 64-channel coil array (3T Siemens MAGNETOM Prisma MR scanner).

### D. Data Analysis

We processed all EEG data in MATLAB (R2022b) including custom scripts, EEGLab (v 2021.0) [47], and BeMoBIL pipeline (v2.0.0) [48] (Fig. 1). We first high-pass filtered raw EEG and EMG data at 1 Hz to remove drift using *eegfiltnew*. We applied a 20-Hz lowpass filter to the EMG channels to remove high frequency noise. We used the EEGLAB plugin *CleanLine* to remove powerline noise at 60 and 120 Hz. Then, we removed channels with a standard deviation 3-times greater than the mean of each subset of data (EEG, noise, and EMG) and performed average referencing [10], [40], [45], [49], [50].

We next used iCanClean to remove noise and muscle artifacts in the cortical EEG electrodes [8], [9], [10]. To perform iCanClean, we used a 4-second moving window with an R^2^ threshold of 0.4 and 0.65 for the EMG and noise channels, respectively [9]. After iCanClean, we did not use the noise and EMG signals for any further analysis. We used the *clean_artifacts* function in EEGLAB to remove noisy cortical channels based on the default parameters, except: chan_crit1 = 0.7, win_crit1 = 0.4, winTol = [-Inf, 10] [10]. Parameters were chosen based on what maximized the number of brain components determined by IClabel while minimizing the number of channels that had to be rejected [10], [51]. EEG data were referenced a final time before we performed adaptive mixture independent component analysis (AMICA) to take the data from individual channels into statistically independent components [52].

We used the MRI of each participant to create custom electrical finite element models with *Fieldtrip* (v. 20210910). We then performed tissue segmentation using *headreco* from the *SimNIBS* toolbox (v 3.2) to segment the scans into six tissue layers (scalp, skull, air, cerebrospinal fluid, gray matter, and white matter) [53]. As described in [10], [14], [40], we generated finite element hexahedral meshes of each participant’s head and co-registered fiducial locations at the left and right helix-tragus junction and nasion from the MRI scan with the structural scan of the EEG electrodes. We then used dipole fitting to model electrocortical activity propagation to the scalp from brain sources using the *<u>ft_dipolefitting</u>* function in the Fieldtrip Toolbox (v20221005). We warped dipole locations to the Montreal Neurological Institute (MNI) template for younger and older adults with Advanced Normalization Tools (ANTs, https://github.com/ANTsX/ANTs) [54]. We identified independent components as brain sources based on a set of established criteria [10]: residual variance < 15%, IClabel identifying the probability of the component being a brain source > 50% [51], negative slope of the power density spectrum from 2-40 Hz, and dipoles located inside the brain. At this stage, we removed an additional ten participants from further analysis because they had less than five brain components and three participants because they had no rest trial. In the remaining participants, we observed 15±5 brain components in younger adults and 12±5 brain components in older adults.

We used a k-means clustering algorithm to determine brain components that were present across participants. Clusters with at least half of the younger adults (n >= 16) and half of the older adults (n >= 30) were retained for group analysis. We established 11 clusters in total. If a participant had multiple components in a cluster, the component with the maximum likelihood to be a brain component according to ICLabel was used and the other component dropped. To further correct for muscle artifact in power spectral density plots (PSDs), we used spectral principal component analysis (sPCA) following the methods and scripts provided in [13], [55]. Details on how sPCA was implemented are provided in Liu, et al., 2025 [41].

For each cluster, we computed the log PSD using *spectopo* from EEGLAB during each task. We then used the Fitting Oscillations & One Over F (FOOOF) toolbox to separate the aperiodic and oscillatory components of the PSD curve [56]. Default parameters were used for separation: range of power spectra set to 3 to 40 Hz; peak width limits of 1 to 8; minimum peak height of 0.05; and maximum number of peaks at 2. The equation to quantify aperiodic EEG was defined as

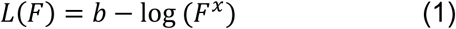

Where b is the broadband offset, x is the exponent, and F is the range of frequency values. A lower value for x indicates flattening of the curve (Fig. 1). The model fit of the extracted aperiodic and periodic components after FOOOF to the original PSD is reported as R^2^. A participants’ aperiodic fit data for a singular brain region was removed if the aperiodic exponent was negative during any of the rest or walking conditions (N = 4 in four different brain areas).

### E. Statistical Analysis

We compared demographic information of waist circumference and subject specific walking speeds between the older and younger adults (MATLAB R2022b). We first used an Anderson-Darling test to check for normality in both datasets and then ran either a t-test or a Wilcoxon rank-sum test if the data was normally or not normally distributed, respectively. We present p-values in the results (α = 0.05).

We performed all additional statistical analyses in R (Version 4.4.3 2025-02-28 UCRT). For our first hypothesis, we use linear mixed effects models with fixed effects for age (young vs. old), task (rest vs. walk), and the interaction of age and task. We further included a random intercept for subject differences. We made separate models for each brain region and the aperiodic exponent and offset. We checked model reliability and normal distributions of residuals using QQ-plots. If the interaction of age and task was not significant, it was removed and the linear model was refit. Post-hoc pairwise comparisons were made only of the fixed effects that were significant (*emmeans* package in R). At most, the following pairwise comparisons were made: younger rest v. younger walk, older rest v. older walk, younger rest v. older rest, and younger walk v. older walk. We corrected for multiple comparisons using the Benjamini-Hochberg procedure to control the false discovery rate. We report the fixed effects estimates (β) and standard errors (SE) from each model alongside p-values (p) from post-hoc tests after there has been adjustment for false discovery rate. If there was an interaction effect in the LME, then both the interaction and post-hoc p-values are reported. We present adjusted p-values in the results (α = 0.05).

For our second hypothesis, we developed a BART model to determine predictors of subject-specific walking speed, measured walking overground on a three-meter walkway, as described in B. Experimental Protocol. BART uses decision trees to model non-linear relationships between a set of predictors and a response variable [36]. Using a Bayesian framework works well with non-parametric data and missing data, determining the best model architecture while preventing overfitting of data [37], [57]. BART has been previously used to understand the biomechanical factors that predict human response to an intervention and to track individual rehabilitation progression [58], [59], [60], [61]. We generated a BART model with the following predictors: participant age, waist circumference, sex, and EEG metrics for the left and right sensorimotor regions. The EEG metrics included: aperiodic offset, aperiodic exponent, average theta power, average alpha power, and average beta power. As a result, the model was generated from a dataset of 90 observations (one for each participant) and 13 predictors. The response variable in the BART model was each person’s subject-specific speed in meters/second (m/s).

We performed hyperparameter tuning using 10-fold cross validation with the *BARTmachineCV* function to generate our BART model (Parameters: *k = 2, q = 0.90, nu = 3, num_trees = 200, seed, = 18*) (*bartMachine* package in R) [57]. We reported pseudo R^2^ and root-mean-squared error (RMSE) as measures of model fit. Given not every participant had data from all brain regions, there was an average of 24% missing data for each model predictor. Thus, the BART model was set to use missing data (*use_missing_data = TRUE*) as a node in the regression trees [57].

We used accumulated local effect (ALE) plots to interpret model outcomes [38]. ALE plots visualize the effect of each model predictor on the response variable while controlling for all other model predictors. We selected ALE plots over other common machine learning visualization techniques, such as partial dependence plots, because they can handle highly correlated data that we expect in a biological dataset [38]. ALE plots do not extrapolate to information not included in the data but only model the relationship between each predictor and the response variable based on the values provided in the data set. We calculated the covariance matrix of our model predictors to aid in interpreting the ALE plots.

We created ALE plots using a bootstrapping procedure of 50 iterations. For each iteration only 75% of the data was used [58]. We binned each predictor’s ALE plot into equally spaced intervals to further smooth the data. ALE plots include lines that connect bins next to each other that have at least one participant in that bin, error bars at each bin indicating one standard deviation from the mean of the 50 iterations of ALE plots, and the data point at the location of each bin is sized to the number of participants in that bin. We then calculated the effect of each predictor on the response variable as the difference between the 90^th^ and 10^th^ percentile of the ALE plot. Thus, the net effect indicates the effect changing that predictor has on the response variable, individualized walking speed, while controlling for all other model predictors.

## III. RESULTS

A total of 90 participants, N = 31 younger and N = 59 older adults, were included in the final analysis (Table I). Older adults walked at significantly slower speeds (p < 0.001) and had significantly larger waist circumferences (p < 0.001).

**Table I.** Participant Characteristics.

|  | Younger | Older | p-value |
| --- | --- | --- | --- |
| Sample Size (N) | 31 | 59 | n.a. |
| Age (years) | $24 \pm 4$ | $74 \pm 6$ | n.a. |
| Sex (M/F) | 15/16 | 23/36 | n.a. |
| Waist Circumference (cm) | $79 \pm 8$ | $97 \pm 13$ | $<0.001$ |
| Individualized Treadmill Speed (m/s) | $0.70 \pm 0.17$ | $0.40 \pm 0.21$ | $< 0.001$ |

### A. EEG Source Analysis and FOOOF Performance

We found eight brain source clusters that were present in more than half of younger and older adults: left presupplementary, right premotor, left and right posterior parietal, left and right sensorimotor, occipital, and mid cingulate. Exact dipole locations can be found in Liu et al., 2025 [41]. The FOOOF model produced excellent fits for both younger and older adults (younger R^2^: 0.99 ± 0.02; older R^2^: 0.99 ± 0.02).

### B. Age Group and Task Comparison

We observed differences in the effect of age group (younger v. older adults) and task (rest v. walk) and their interaction on aperiodic exponent and offset in different brain areas (Fig. 2). In the right premotor area, we observed a significant effect of age on both exponent (β = −0.221, SE = 0.068, p = 0.004) and offset (β = −0.457, SE = 0.159, p = 0.006). Older adults had lower exponents and offsets. We also observed a significant effect of task on aperiodic exponent in the right premotor area (β = 0.058, SE = 0.024, p = 0.020), with higher exponents during walking. In the left presupplementary motor area, we observed a significant effect of task on both the exponent (β = 0.067, SE = 0.023, p = 0.005) and offset (β = 0.067, SE = 0.030, p = 0.029), with higher exponents and offsets during walking compared to rest. There was no effect of age for either the exponent or offset in the left presupplementary motor area.

**Fig. 2.**
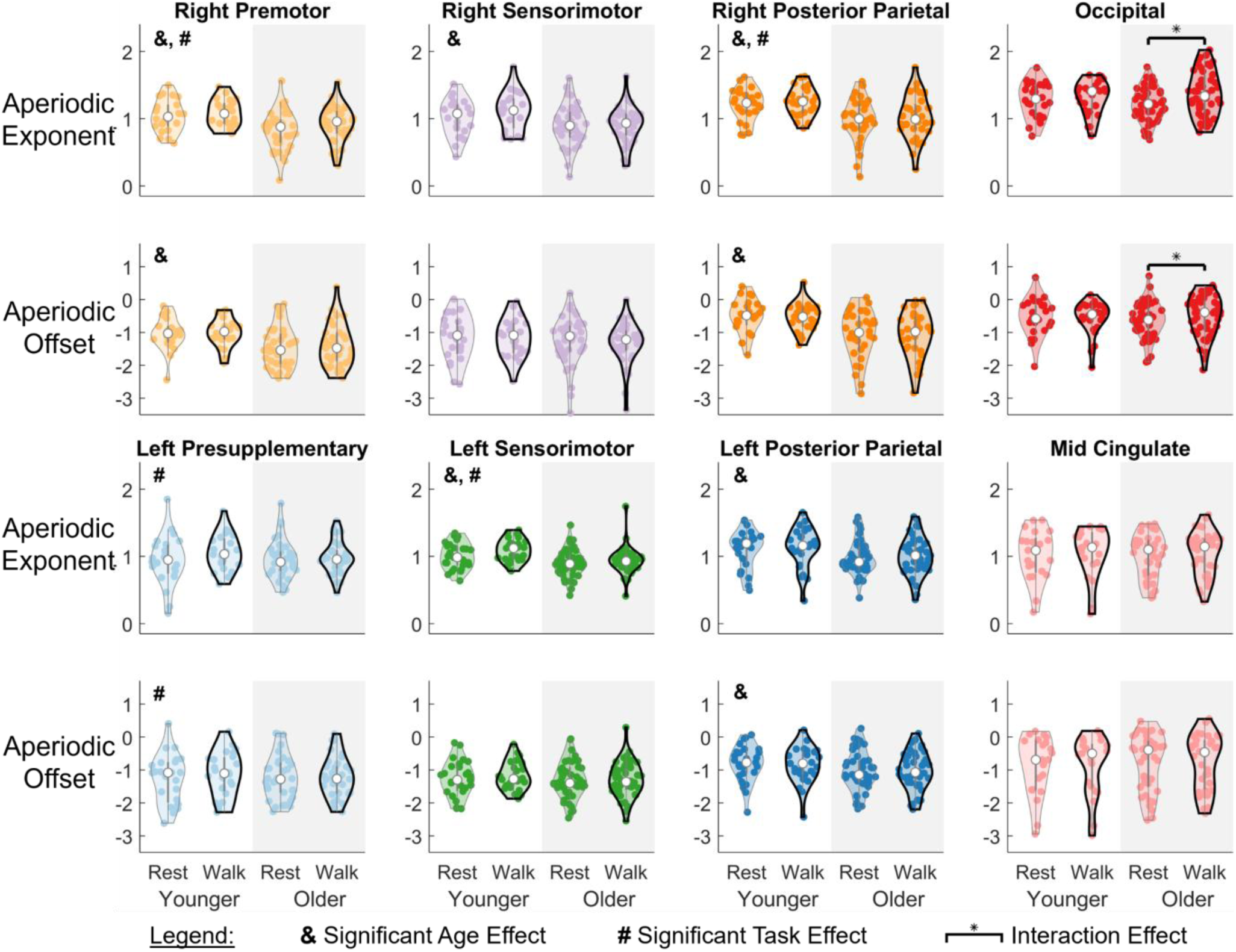
Violin plots comparing the aperiodic exponent and offset during seated rest and walking on a treadmill at a preferred speed in older and younger adults. Asterisks indicate adjusted p-value less than 0.05. Within each plot, the violin plots with a light grey outline are during rest and the bold black outline are during walking, while the plots with a white background are the younger adults and the grey background are older adults. Differences in aperiodic exponent and offset in younger and older adults at rest and during walking. The presence of age-related differences in aperiodic EEG differed across brain regions. Only in the occipital lobe was there a significant interaction between task and age. & indicates a significant effect of age while # indicates a significant effect of task, as shown in the upper left corner of each plot. Significant interactions are shown with an asterisk and a bar between significantly different groups.

In both sensorimotor areas, there were significant age differences in the aperiodic exponent but not the offset. In the right sensorimotor region, we only observed a significant effect of age (β = −0.264, SE = 0.086, p = 0.003), but not task, on aperiodic exponent. Older adults had a lower aperiodic exponent than younger adults. In the left sensorimotor area, there was a significant effect of age (β = -.183, SE = 0.073, p = 0.015) and task (β = 0.068, SE = 0.021, p = 0.003) on the exponent, but no significant effect of the interaction of age and task. Older adults had significantly lower exponents in the left sensorimotor area, and aperiodic exponent was higher during walking compared to rest.

We observed an age effect in both the left and right posterior parietal areas on the aperiodic exponent (right: β = −0.329, SE = 0.104, p = 0.005; left: β = −0.169, SE = 0.066, p = 0.012) and aperiodic offset (right: β = −0.709, SE = 0.182, p < 0.001; left: β = −0.301, SE = 0.141, p = 0.036). Older adults had lower aperiodic offset and exponents in both the left and right sensorimotor areas. Aperiodic exponent was also higher during walking in the right posterior parietal area (β = 0.057, SE = 0.024, p = 0.023).

Only in the occipital area did we find interactions between age and task for both the aperiodic exponent (β = 0.132, SE = 0.063, interaction p = 0.038) and offset (β = 0.227, SE = 0.093, interaction p = 0.018). Post-hoc tests indicated older adults had significantly higher aperiodic exponents (p < 0.001) and offsets (p = 0.002) during walking compared to rest, but younger adults had no difference between rest and walk. There were no significant effects of age, task, or their interaction in either the aperiodic exponent or offset in the mid cingulate.

### C. Aperiodic Fit as a Predictor of Walking Speed

The BART model (R^2^ = 0.70, RMSE = 0.13 m/s) identified which predictors (numerical age, sex, waist circumference, aperiodic offset, aperiodic exponent, average alpha power, average beta power, or average theta power in the left and right sensorimotor areas) had the largest effect on walking speed. As expected, some model predictors were moderately correlated, with the highest correlations between aperiodic offset and exponent within the same brain region (Supplemental Fig. 1).

Among all model predictors, waist circumference, age, and sex had the largest net effects on walking speed at 0.22 m/s, 0.18 m/s, and 0.15 m/s, respectively (Fig. 3 and Fig. 4). Older participants and those with a larger waist circumference walked at slower speeds, while men walked faster than women. Right sensorimotor alpha had the next largest net effect on walking speed at 0.11 m/s, with a higher right sensorimotor alpha for those with faster walking speeds. Both the left sensorimotor offset and beta had a net effect of 0.07 m/s and both were lower for those with faster walking speeds (Fig. 5). The right sensorimotor offset (net effect = 0.06 m/s) was also lower for those with faster walking speeds, while both right and left sensorimotor exponents were higher for those with faster speeds (net effect = 0.05 m/s and 0.04 m/s, respectively).

**Fig. 3.**
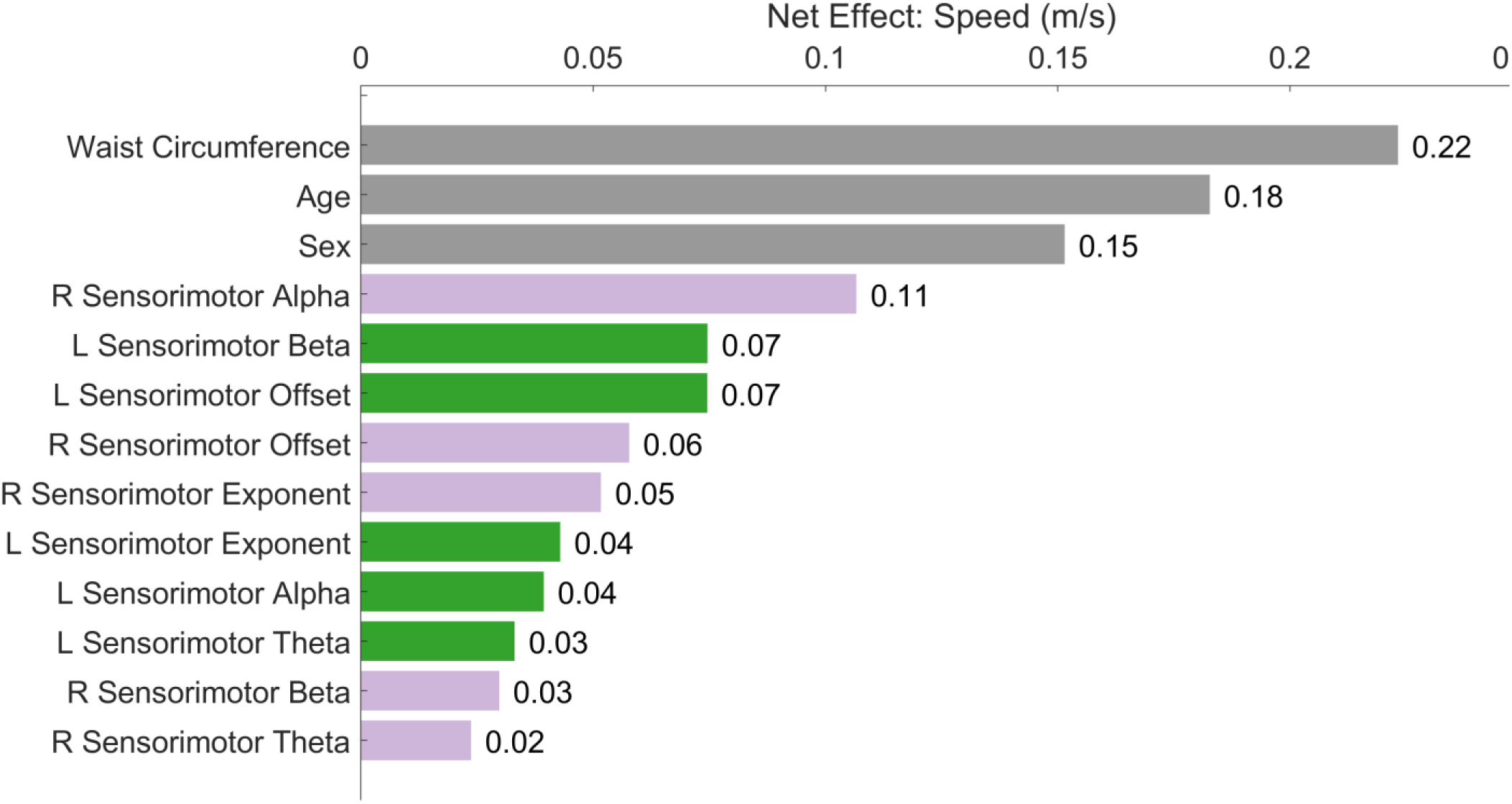
Effect size of each parameter on walking speed in meters/second (m/s) when controlling for all other model predictors. L and R denote left and right sides, respectively. Waist circumference, age, then sex had the largest effect on walking speed, followed by the right sensorimotor alpha and the left sensorimotor beta and offset.

**Fig. 4.**
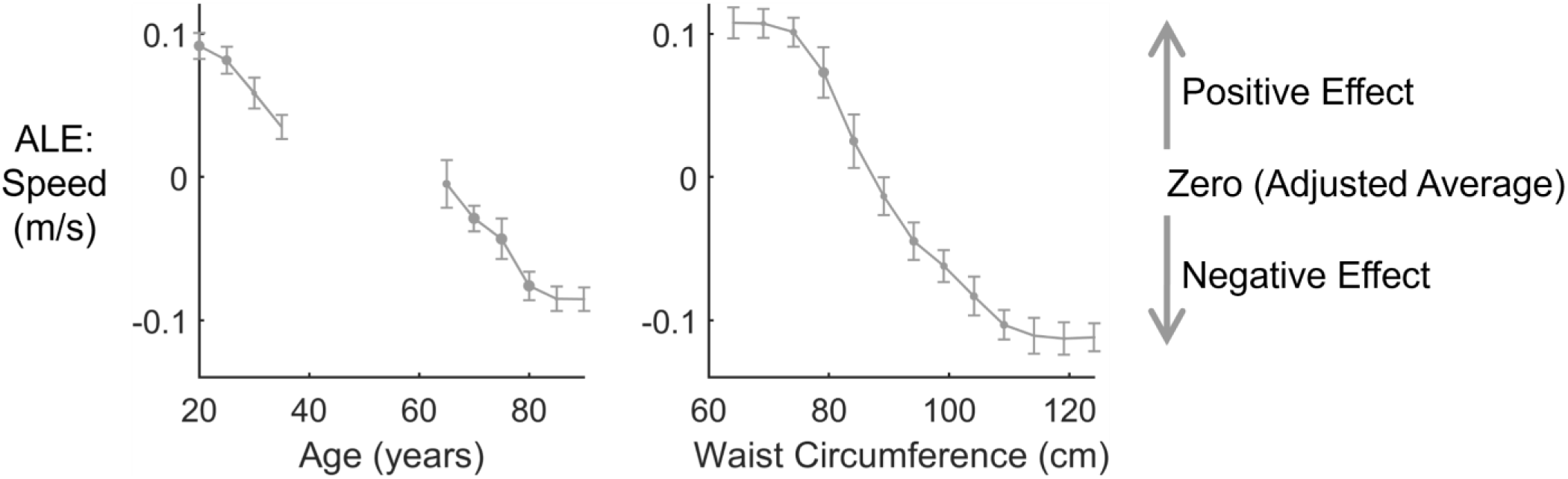
The accumulated local effects (ALE) plots of age and waist circumference in centimeters (cm). ALE plots demonstrate the effect of each model predictor on the response variable, individualized walking speed, while controlling for all other model predictors. A positive number indicates a faster speed from the adjusted average after controlling for all other model predictors, while a negative number represents a lower speed relative to the adjusted mean. Vertical bars indicate ±1 standard deviation form 50 ALE plot iterations and the size for each point represents the number of subjects in that bin. Individualized walking speed deceased while and age and waist circumference increased.

**Fig. 5.**
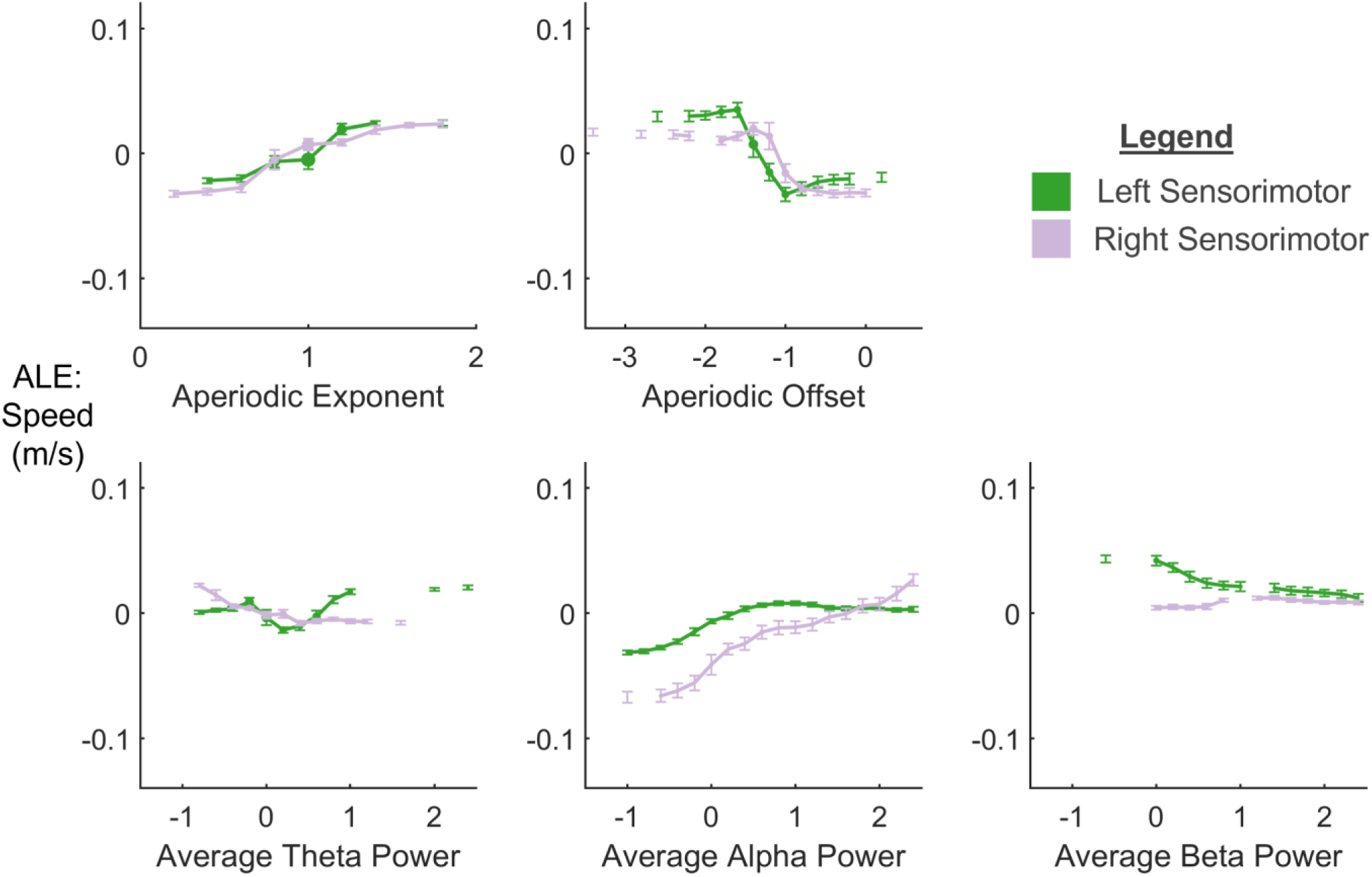
ALE plots of the EEG outcomes (aperiodic exponent, aperiodic offset, average theta power, average alpha power, and average beta power) for the left and right sensorimotor area. Vertical bars indicate ±1 standard deviation form 50 ALE plot iterations and the size for each point represents the number of subjects in that bin. Aperiodic exponent was higher at faster speeds, while aperiodic offset was lower at faster speeds for both left and right sensorimotor areas. Right and left sensorimotor alpha power were higher at faster speeds, while left sensorimotor beta was lower at faster speeds. Most other oscillatory EEG power varied little at higher individualized walking speeds.

The remaining brain region aperiodic parameters had a net effect at or below 0.04 m/s (Fig. 3). These parameters included the right sensorimotor beta and theta and left sensorimotor alpha and theta. Right sensorimotor theta was lower for those with faster walking speeds, while left sensorimotor alpha, right sensorimotor beta, and left sensorimotor theta had a non-monotonic relationship with participant walking speed.

## IV. DISCUSSION

We found that age differences in aperiodic EEG were present during rest and walking but differed between brain areas. Previous studies have only reported aperiodic EEG acquired with participants in a seated position and without source localization to quantify brain area-specific aperiodic EEG. Our results suggest age-specific effects on the underlying physiological mechanisms driving aperiodic EEG in different brain areas. We also quantified which EEG metrics in the left and right sensorimotor regions were the strongest predictors of walking speed. We found that right sensorimotor alpha power was the strongest predictor of walking speed, followed by left sensorimotor beta and offset, after controlling for age, waist circumference, and sex. Quantifying both aperiodic and oscillatory EEG in future studies will give a more comprehensive understanding of brain function.

Our findings support prior research that aperiodic exponent and offset are lower in older adults compared to younger adults [17], [24], [25], [26], [56]. We observed a lower aperiodic exponent in five of eight brain regions, and lower offset in three of eight brain regions in older adults compared to younger adults. These findings suggest that age-related changes in aperiodic EEG are not present in all brain areas but are present both at rest and while walking. No other studies have reported aperiodic EEG during a movement task or after performing source localization, but age-related shifts in aperiodic EEG have been shown during cognitive tasks performed at a seated rest [16]. The age group differences in aperiodic EEG persisting between tasks supports the evidence that aperiodic EEG may be an indicator of brain health in clinical populations [29]. Even with this consistency across tasks, aperiodic EEG characteristics can differ across electrode channels [25], [30]. Prior work at the channel level indicates that the correlation between aperiodic EEG and age of older and younger adults differs across EEG channels [24], [30], and changes with development in children [62]. Our findings suggest that age-related shifts in aperiodic EEG may be driven by only a subset of brain areas. Signals from one brain region can propagate to many EEG channels around the head [63]. One EEG channel capturing electrical activity in several brain regions likely explains why Donoghue et al., 2020 reports significantly lower aperiodic exponent and offset in older adults from just a single electrode over the occipital lobe while we saw no main effect of age in the occipital lobe after source localization [56]. Changes in aperiodic EEG in some brain areas may be large enough to drive significant differences in aperiodic exponent and offset across several EEG channels. We also observed that the left and right sensorimotor exponents, but not offsets, were significantly lower in older adults. Thus, the underlying mechanisms contributing to aperiodic exponent and offset in the sensorimotor areas may differ in how they are affected by aging.

The left presupplementary, left sensorimotor, right premotor, and right posterior parietal areas all had higher aperiodic exponents (i.e. less flattened curve) during walking compared to rest. One possible explanation for higher exponents between tasks is that the brain reduces excitation or increases inhibition in these areas during walking [22], [23], [64], [65], [66]. Future research is needed to determine the exact physiological mechanisms driving exponent changes between tasks. However, modulation of the aperiodic exponent in these areas of the brain during different tasks would suggest they are being relied on differently during seated rest and walking. Only in the left presupplementary cortex did offset increase while walking compared to rest. Increased neuronal firing rates could be driving increased aperiodic offset while walking, suggesting the left presupplementary area is more active during walking [20], [21].

We only observed a significant interaction between age and task in the occipital lobe. Older adults had a higher exponent and offset during walking compared to rest while younger adults did not. Previous work has shown that older adults rely more on visual information for sensory feedback while walking than younger adults [67], [68]. This greater reliance on vision may alter aperiodic EEG in older adults during walking compared to sitting. It is also possible that the increase in exponent and offset in the occipital lobe represents neural compensation for other brain regions [69]. While aperiodic exponent and offset increased during walking in some brain areas, it was never enough to mask the age differences.

It is important to note that physiological variables other than cortical activity may contribute to aperiodic EEG if signal cleaning is not able to fully remove them. For example, heart rate variability has been shown to correlate with the lower aperiodic EEG observed in older adults [70]. However, we have performed additional cleaning than this prior work that may reduce the influence of other physiological signals on EEG, such as using iCanClean to remove EMG data and performing source localization. Additionally, we observed differences between aperiodic EEG in the different brain regions. The differences in aperiodic EEG are not uniform across brain areas, suggesting that an underlying biological signal visible across the brain, such as heart rate, is not the only signal contributing to aperiodic EEG.

We found that age, waist circumference, and sex had the largest effect on walking speed in our BART model. This finding was not surprising given that walking speed is known to be lower in older adults, people with a larger waist circumference, and in women compared to men [71], [72], [73], [74]. By including these demographics in the BART model, we were able to control for their overall effect on walking speed to improve the accuracy of detecting the effect of other model predictors. For example, we know older adults walk at slower speeds, but aperiodic exponent is still higher in people with faster walking speeds while aperiodic offset is lower in people with faster walking speeds after controlling for age in the left and right sensorimotor areas. This relationship between offset, speed, and age, suggests that decreasing aperiodic offset could reflect a compensatory mechanism for reduced mobility function in older adults. However, future work will need to be done to assess possible neural compensation and its effects on walking speed. In addition, previous work found that older adults grouped by their slower and faster preferred speeds did not have significant differences in their alpha power [14]. But after controlling for age, waist circumference, sex, and other EEG metrics, right sensorimotor alpha was higher in those with a higher individualized walking speed. It is understood that different EEG metrics can influence each other [29], so using the BART model with ALE plots may reveal relationships not apparent in group statistical analysis by controlling for other variables included in the model.

The minimum clinically important difference (MCID) for walking speed represents how much improvement in walking speed results in a meaningful outcome in daily life. The MCID of walking speed for older adults with a pathology is 0.1 m/s, comparable to some of the net effects of EEG metrics on walking speed in this study [75], [76]. It is unknown if the EEG metrics can be adjusted with intervention or if this would have a direct effect on walking speed, but this effect may be something to consider in future studies. Prior work in other populations indicates that certain interventions, such as brain stimulation, may be able to alter aperiodic and oscillatory EEG signal [29], [77], but future work needs to consider mobility in older adults.

This study presents novel results on how aperiodic EEG is related to slower walking in older adults. However, there are several limitations that should be considered in the evaluation of this work. First, the brain areas included in the analysis are based on what sources were present in at least half of younger adults and half of older adults. Thus, the findings are not representative of every brain region’s aperiodic EEG. This study also had strict inclusion and exclusion criteria to focus the study population on healthy adults. As a result, the range of individualized treadmill speeds presented here may not represent older adults who walk at slower speeds due to poorer health status. Additionally, this study is unable to give insight into what exact underlying neurological mechanisms are responsible for the observed differences in aperiodic EEG. However, these findings in older and younger adults motivate a need to better understand the physiological drivers of aperiodic EEG, and whether they can explain mobility decline in older adults. There are also some limitations to consider for the BART model specifically. The individualized treadmill speed used as the BART outcome was set based on the speed in which someone could comfortably walk on uneven terrain, not purely preferred overground walking speed. Thus, the net effects may vary slightly if true preferred walking speed were used. Adding additional model predictors, such as other brain regions, could likely improve model fit. However, prior work indicates that the ALE plots remain accurate, even with a lower R^2^ value due to high variability in the dataset not captured by a BART model [61]. We aimed to maintain a lower number of model predictors relative to our number to subjects based on a priori hypotheses about the importance of sensorimotor areas involved in walking. Future work could include larger sample sizes, or alternative methods to reduce the number of model predictors prior to building the model [78], [79], [80].

## V. CONCLUSION

This is the first study to quantify aperiodic EEG in specific brain regions during walking in older and younger adults. We found that older adults only had lower aperiodic EEG in some brain areas, but these age-related differences were present during both rest and walking tasks. We also found that walking increases aperiodic EEG compared to rest in a subset of brain areas. We further applied machine learning techniques to model the non-linear and highly variable relationship between walking speed and several model predictors: age, waist circumference, sex, and EEG metrics in the sensorimotor brain regions. We found that right sensorimotor alpha power, left sensorimotor offset, and left sensorimotor beta had the largest effect on walking speed compared to other aperiodic and oscillatory EEG outcomes in the sensorimotor areas. These results suggest that both oscillatory and aperiodic EEG may give insight into functional. Future studies should consider both aperiodic and oscillatory EEG in their analyses and determine if there is the possibility to target these EEG metrics with interventions supporting mobility in older adults.

## Data Availability

All data produced in the present study are available upon reasonable request to the authors.

