## Supplemental Figure 1 for "Mobility Function and Aperiodic Electrocortical Activity in Younger and Older Adults"

*Supplemental Figures*

Charlotte R. DeVol, Chang Liu, Jacob Salminen, Erika M. Pliner, Arkaprava Roy, Chris J. Hass, David J. Clark, Todd M. Manini, Rachael D. Seidler, Daniel P. Ferris

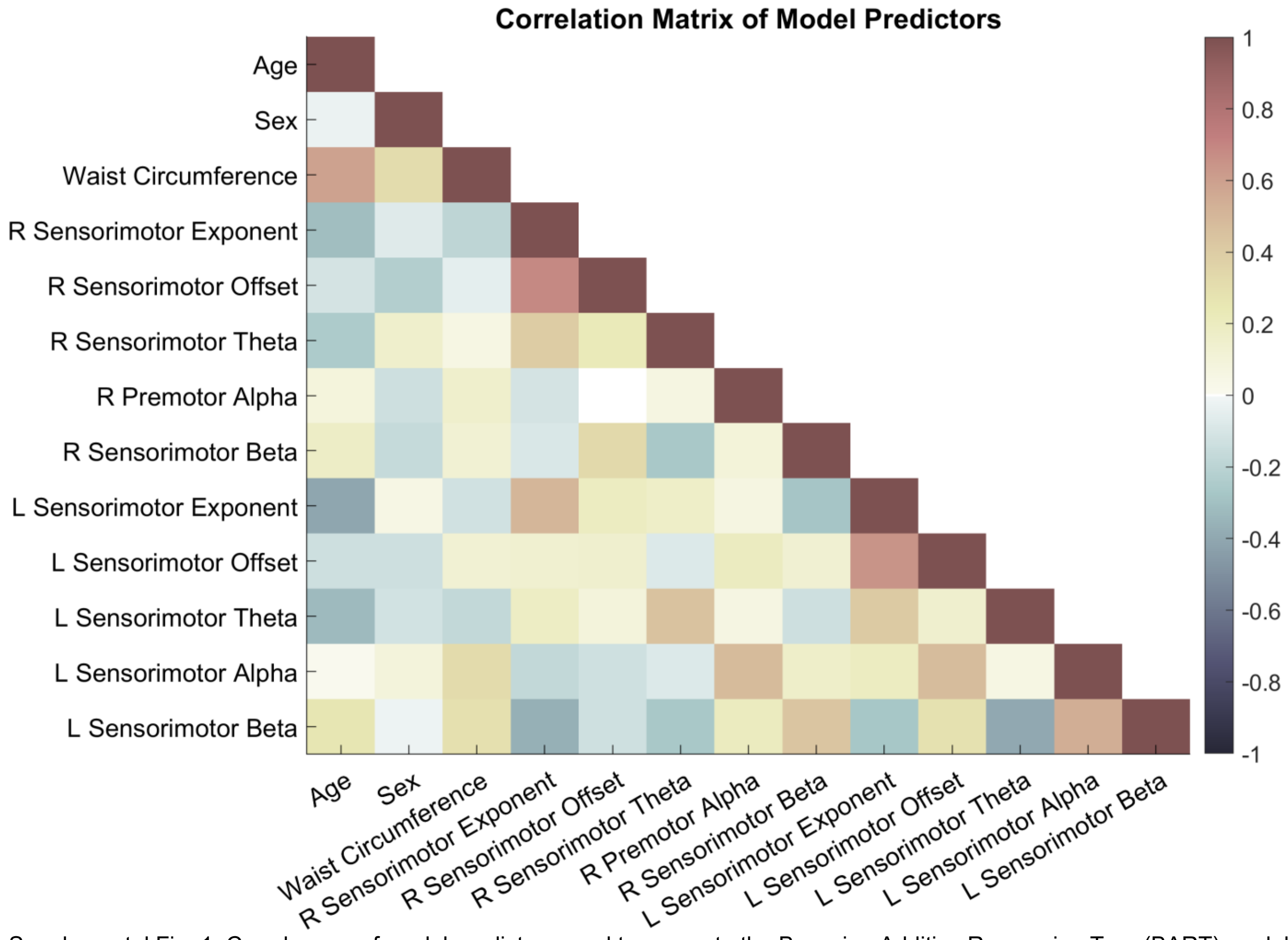


Supplemental Fig. 1. Correlogram of model predictors used to generate the Bayesian Additive Regression Tree (BART) model. The correlogram is given to aid in interpreting ALE plots. ALE plots only represent the relationship between each BART model predictor and the response variable (individualized walking speeds) given the relationships that exist in the data set. The strongest correlations were seen between the aperiodic offset and exponent for each brain region.
